# “It affects every aspect of your life”: A qualitative study of the impact of delaying surgery during COVID-19

**DOI:** 10.1101/2022.01.20.21267627

**Authors:** KM Sauro, C Smith, J Kersen, E Schalm, N Jaworska, P Roach, S Beesoon, ME Brindle

## Abstract

**Background:** The COVID-19 pandemic has overwhelmed healthcare systems, leading many jurisdictions to reduce surgical services to create capacity (beds and staff) to care for the surge of patients with COVID-19. These decisions were made in haste, and little is known about the impact on patients whose surgery was delayed. This study explores the impact of delaying non-urgent surgeries on patients, from their perspective.

**Methods:** Using an interpretative description approach, we conducted interviews with adult patients and their caregivers who had their surgery delayed or cancelled during the COVID-19 pandemic in Alberta, Canada. Trained interviewers conducted semi-structured interviews. Interviews were iteratively analyzed by two independent reviewers using an inductive approach to thematic content analysis to understand key elements of the patient experience.

**Results:** We conducted 16 interviews with participants ranging from 27 to 75 years of age with a variety of surgical procedures delayed. We identified four interconnected themes: individual-level impacts (physical health, mental health, family and friends, work, quality of life), system-level factors (healthcare resources, communication, perceived accountability/responsibility), unique issues related to COVID-19, and uncertainty.

**Interpretation:** The patient-reported impact of having a surgery delayed during the COVID-19 pandemic was diffuse and consequential. While the decision to delay non-urgent surgeries was made to manage the strain on healthcare systems, our study illustrates the consequences of these decisions. We advocate for the development and adoption of strategies to mitigate the burden of distress that waiting for surgery during and after COVID-19 has on patients and their family/caregivers.

## Introduction

The novel SARS-CoV-2 virus (COVID-19) was declared a pandemic by the World Health Organization (WHO) in March 2020,^(1)^ and healthcare systems across the globe braced for a potentially large influx of COVID-19 patients within hospitals. These situations played out in countries such as Italy, where healthcare systems quickly became overwhelmed.^(2)^ In light of the impact of the COVID-19 pandemic on healthcare systems globally, many Canadian provinces reallocated healthcare resources to care for COVID-19 patients by reducing surgical capacity. Consequently, a staggering number of non-urgent surgeries (surgeries not immediately threatening life or limb)^(3)^ were delayed. For example, early in the COVID-19 pandemic response, Ontario delayed 185,000 surgeries, while the number of delayed surgeries during the fourth wave in Alberta climbed to over 30,000 – a number that continues to increase at the time of writing.^(1, 4-6)^ The impact of delaying non-urgent surgeries in Canada has not been fully explored, but it is estimated that the backlog from just the first wave of COVID-19 in some provinces will take 84 weeks to clear.^(6)^

Pre-pandemic evidence suggests that excessive surgical wait times can lead to poor physical health, increased anxiety, decreased social interaction and ability to work and overall quality of life.^(7, 8)^ Factors that mediate the impact of delays in access to surgical care include patient choice in the delay and communication from healthcare providers.^(7, 8)^ It is unclear if these pre-pandemic factors are consistent with the effects of delaying surgery in the context of the COVID-19 pandemic because evidence of the impact of delayed surgeries during COVID-19 is still in its infancy.^(9)^ With the continued rise in delayed surgeries understanding the scope of the personal impact on patients and their daily lives is of critical importance. To address this knowledge gap, the objective of this study is to understand the patient’s perspective of having a surgery delayed during the COVID-19 pandemic response.

## Methods

We adopted an interpretative descriptive approach as our methodological framework, which is aligned with the constructivist and naturalistic orientation of inquiry and aims to understand a phenomenon grounded in the data within a clinic context to apply the findings.^(10-13)^

### Participants

We included patients and family/caregivers of patients in Alberta who had their surgery delayed due to the COVID-19 pandemic response. To date, the Province of Alberta has experienced four waves of COVID-19; the first wave (March 13, 2020 - early May 2020) resulted in a reported 30% decrease in surgical capacity, the second and third wave had no strategic decrease in surgical capacity, and the fourth wave (September 2021 to the time of writing in November 2021) resulted in all non-urgent surgeries being delayed (only emergent and urgent surgeries performed) with an estimated 60-70% decrease in surgical capacity.

### Recruitment and sample selection

Recruitment posters were distributed through social media (Twitter, Facebook, Instagram) and local news outlets to allow us to reach patients across broad networks spanning the province of Alberta. Patients with delayed surgeries and their family/caregivers contacted the Principal Investigator (KMS) via email or phone.

Consistent with interpretative descriptive approach, we used a theoretical sampling strategy – we continued collecting data to achieve a purposive sample of anticipated variations in response based on age, gender, type of surgery and geographical location.^(11)^

### Data collection

Ten interviews were anticipated but we continued until we reached saturation (we no longer identify new themes or novel data) and recruited patients with varied ages, gender, type of surgery and geographical location.

Interviews were conducted by experienced facilitators (CS, JK, ES, NJ). The facilitators were female graduate students and a research associate with experience in qualitative methods, but not within the area of surgical care or COVID-19, who volunteered to conduct the interviews. Therefore, there were few assumptions or bias with regards to the results of the study. All facilitators had formal graduate training in qualitative methods or experiential training in interview facilitation. The Principal Investigator and facilitators did not have previous relationships with the participants or their healthcare providers.

The experienced facilitator used a semi-structured interview guide with prompts to elicit emergent ideas while maintaining focus on the topic (Appendix E – Interview Guide). The interview guide was pilot tested amongst the research team and feedback was solicited from the senior researcher (MEB) who is a surgeon and leader within the healthcare system.

Interviews were conducted virtually (using Zoom® or telephone) due to public health restrictions in Alberta during data collection, were audio recorded and transcribed. Field notes were included to clarify context, where appropriate.

### Data analysis

Data were managed using NVivo 12.0^(14)^ and analyzed in keeping with our interpretive descriptive approach, by two independent reviewers using thematic content analysis to understand key elements of the patient experience.^(11-13)^ An inductive approach was used so themes were drawn from the data rather than pre-existing sources. Data analysis was iterative whereby the two reviewers coded transcripts, met to compare emerging themes to ensure rigour of the analytic process. Final themes were determined by consensus among reviewers. The results were reviewed by the interview facilitators and three participants to ensure trustworthiness of the findings.

This study was approved by the University of Calgary Conjoint Research Ethics Board (REB20-0753).

## Results

We conducted 16 interviews between September 20, 2021 and October 8, 2021. The average length of the interviews was 22 minutes. Demographic characteristics of participants are described in Table 1. Briefly, the mean age of the participants was 47 years old, with the majority identifying as women. Most surgeries were scheduled in urban centres and the type of scheduled surgery varied considerably. Four participants had their surgery completed prior to the interview while nine did not.

**Table 1.**
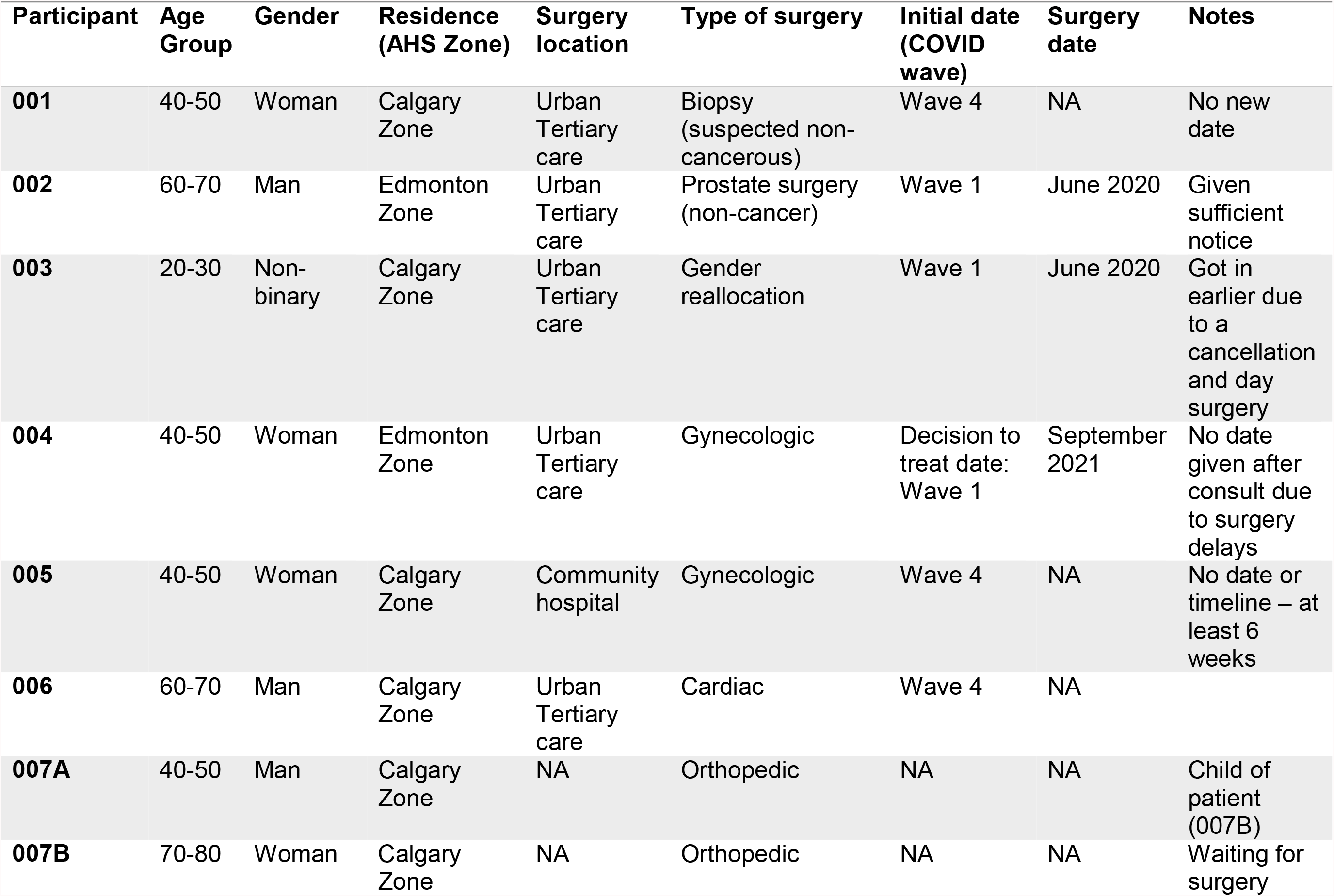
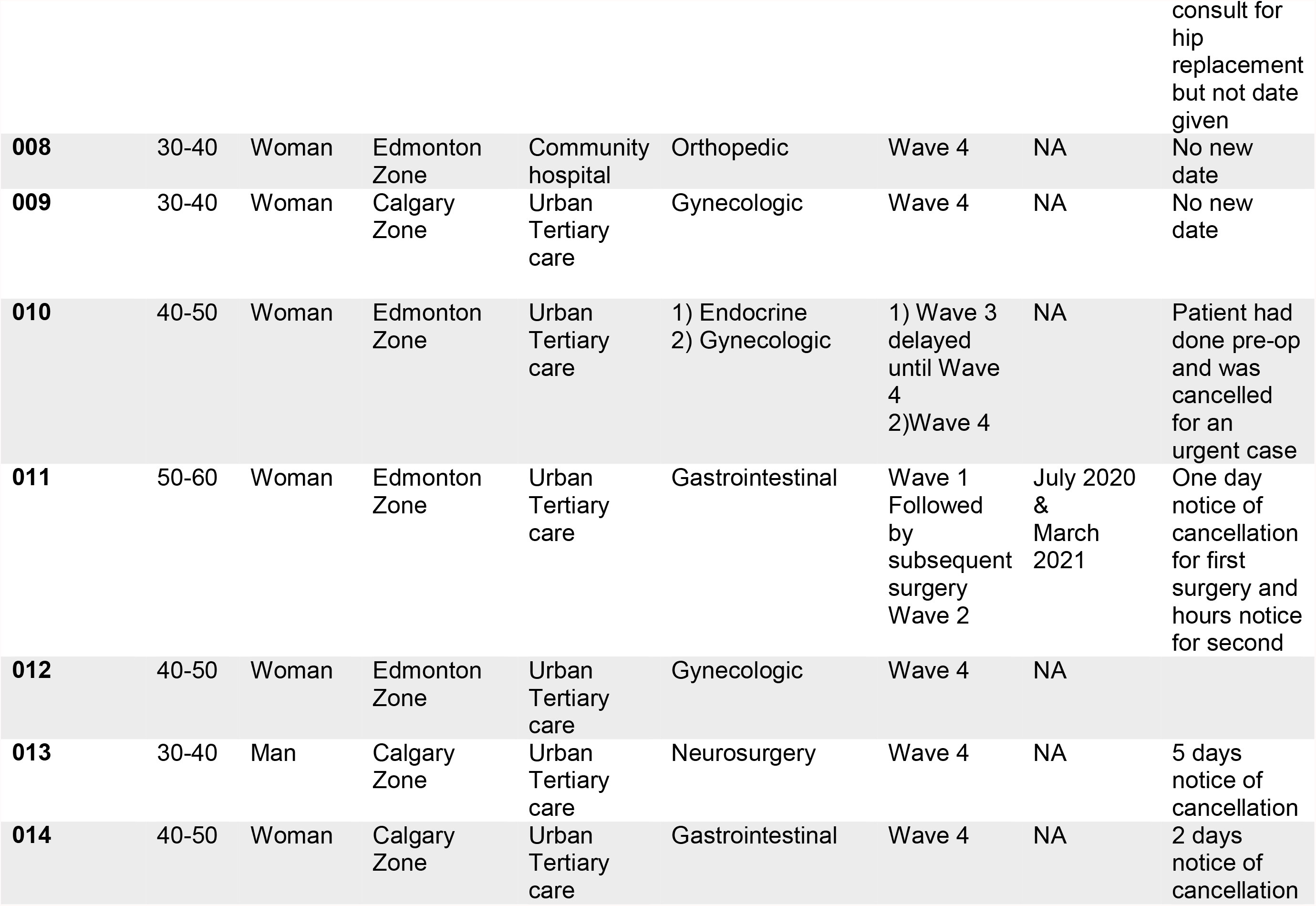
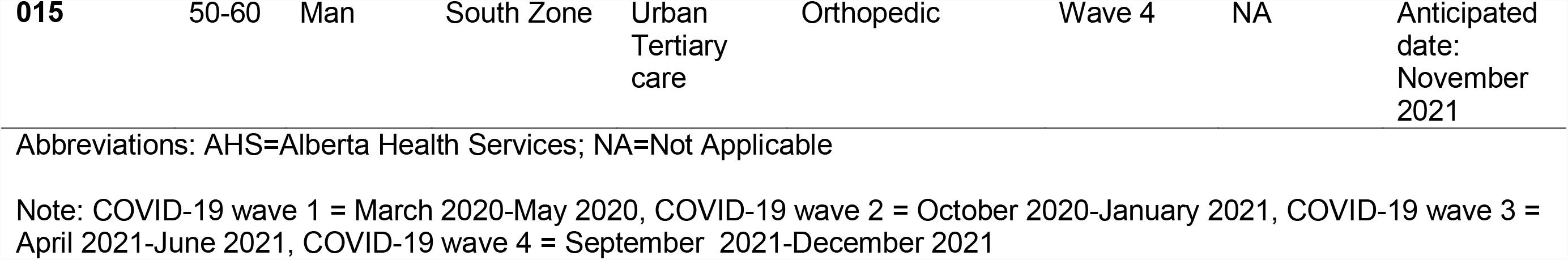
Participant characteristics

### Themes

There were four overarching themes that emerged from the data (Figure 1): 1] individual-level impacts (subthemes: physical health, mental health, family and friend impact, work impact, quality of life), 2] system-level factors (subthemes: communication, healthcare resources, perceived accountability/responsibility), 3] COVID-19-related impacts and 4] uncertainty. Table 2 describes each theme and provides example quotes.

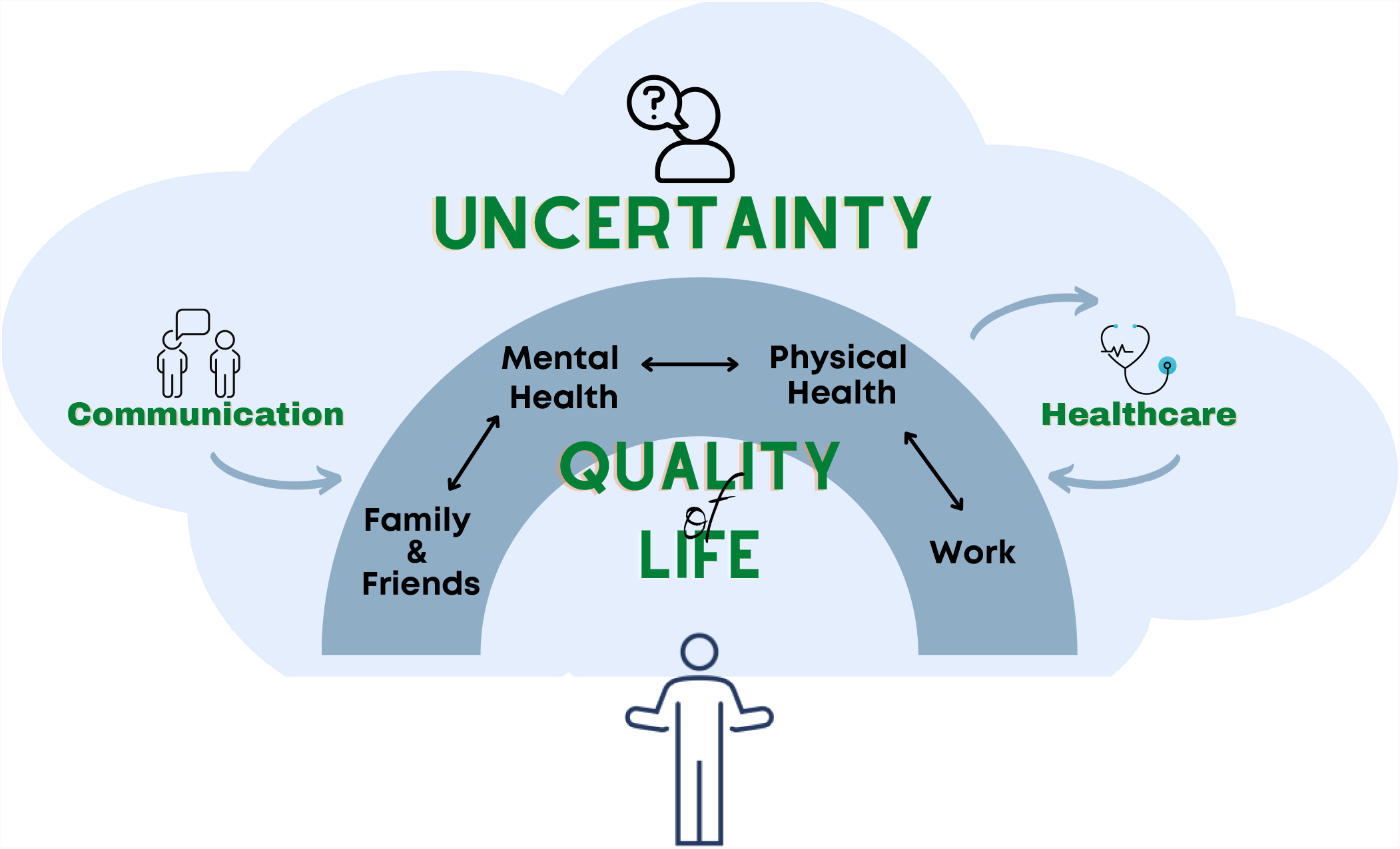

**Table 2.**
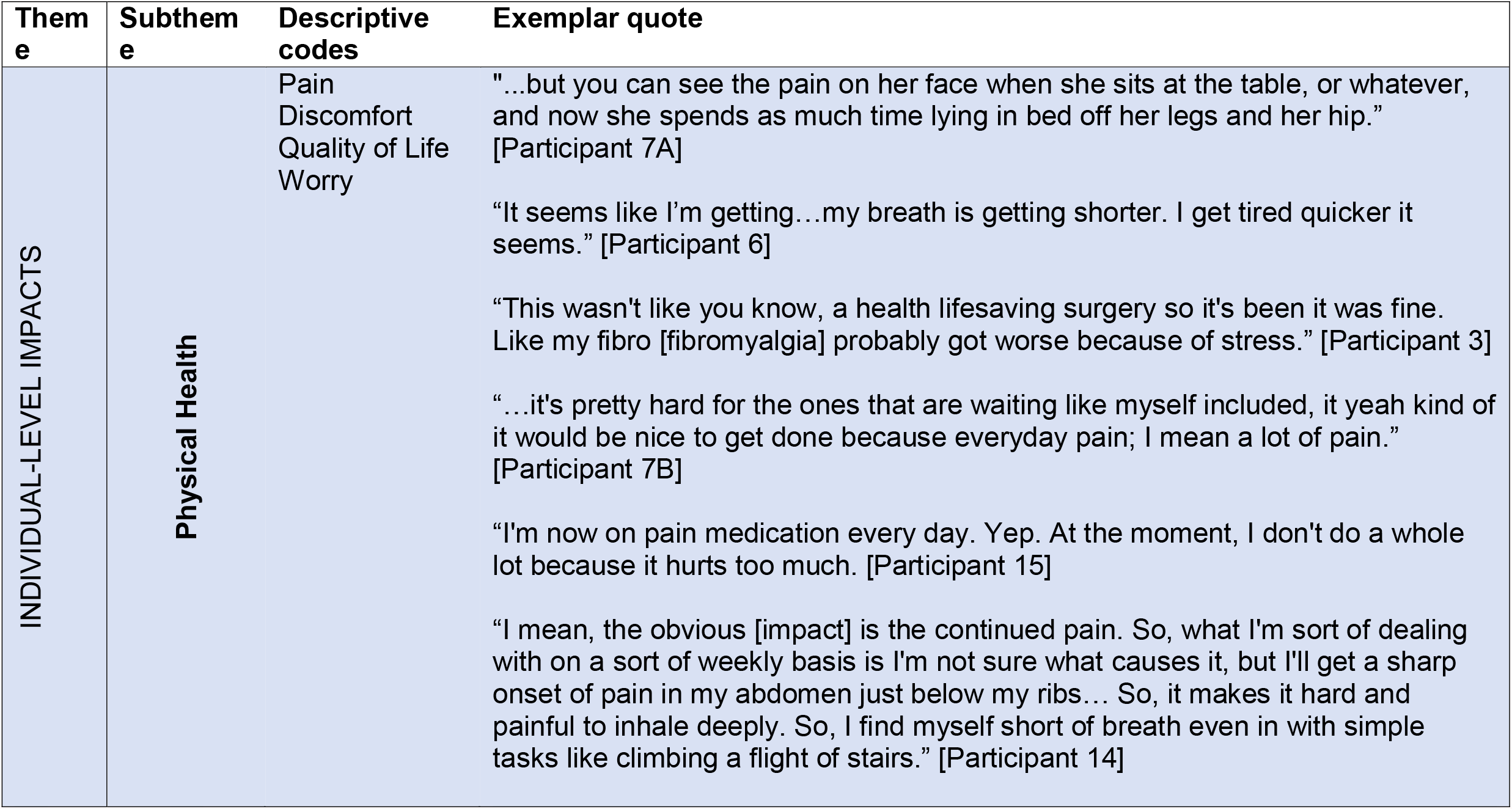
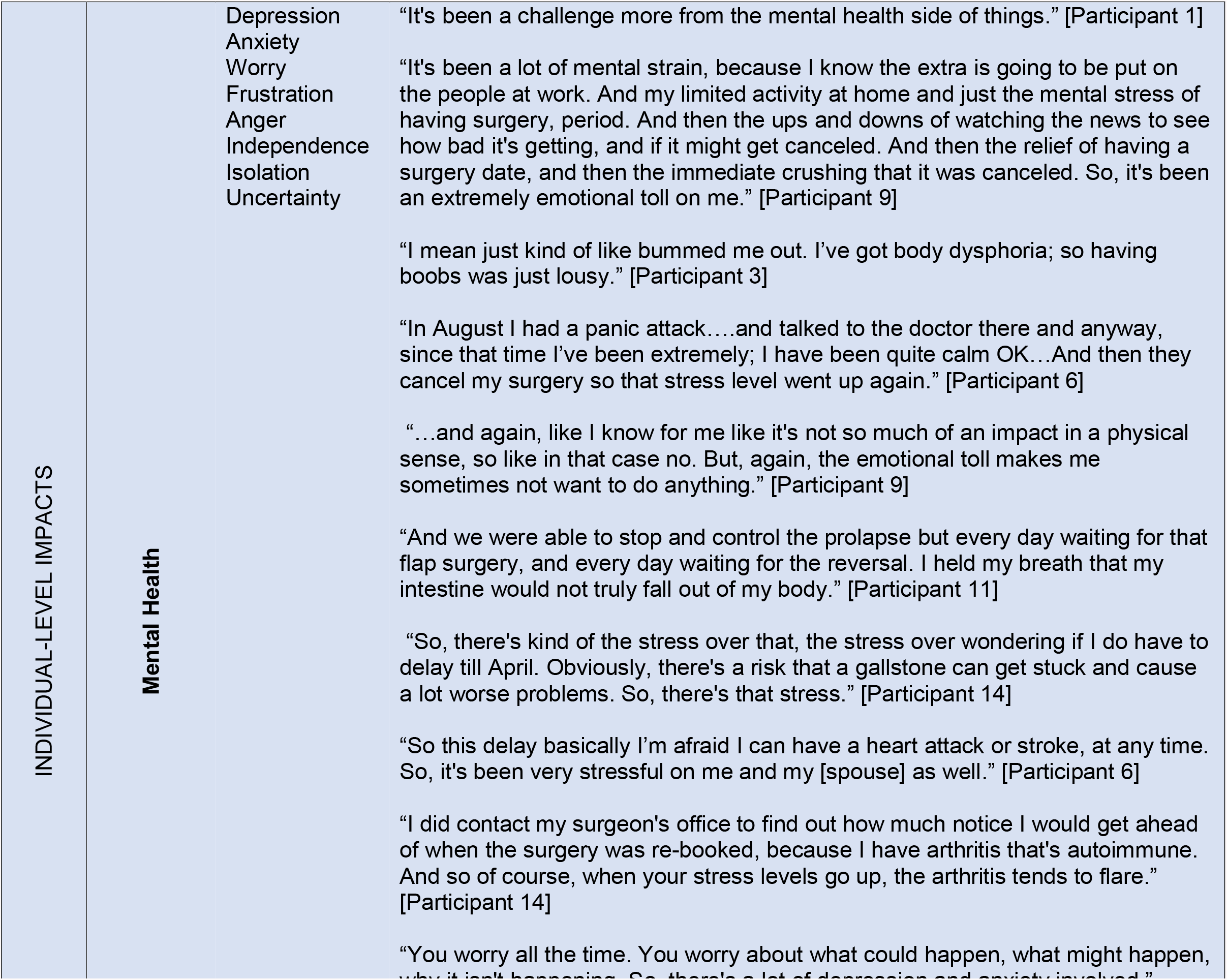
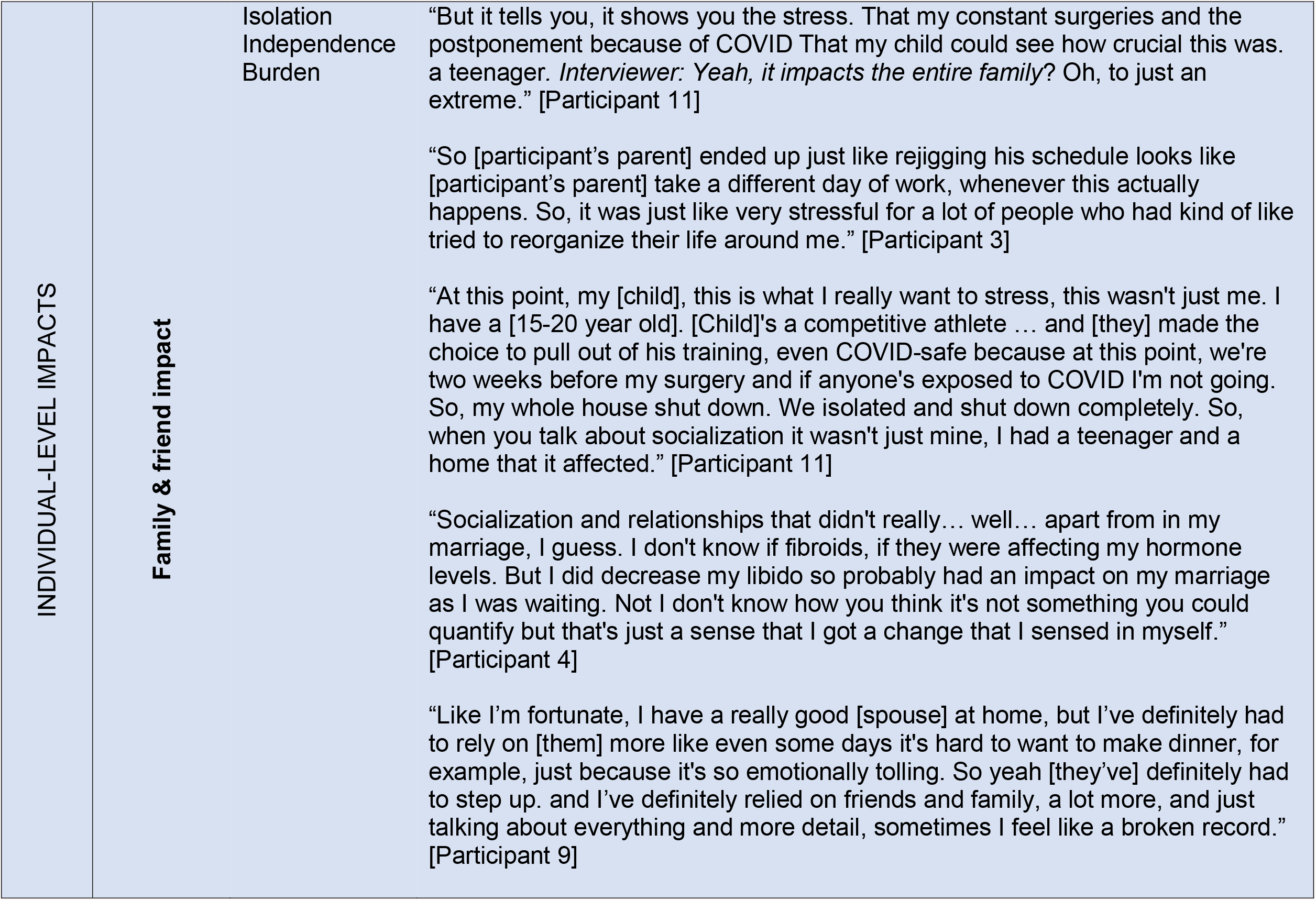
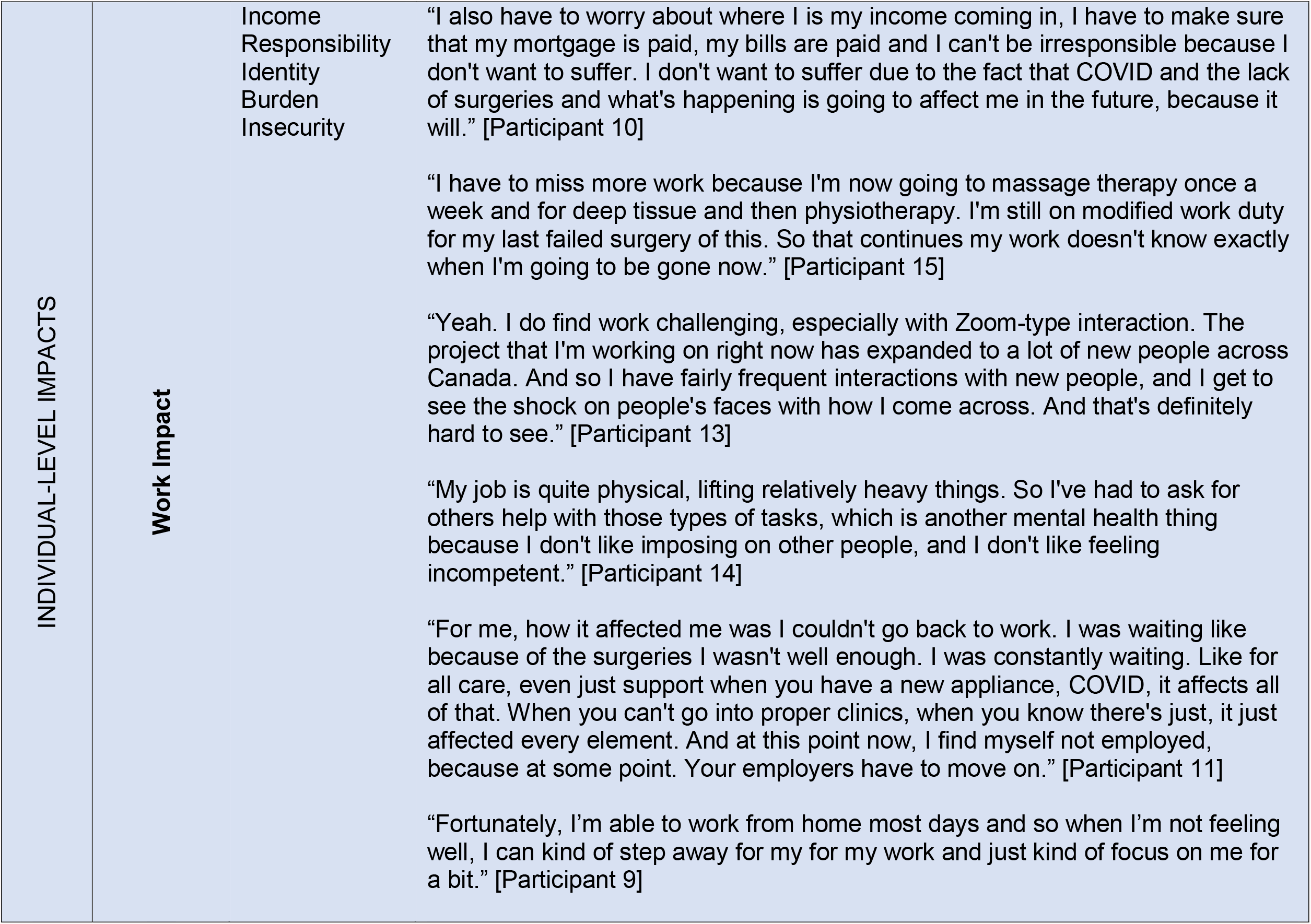
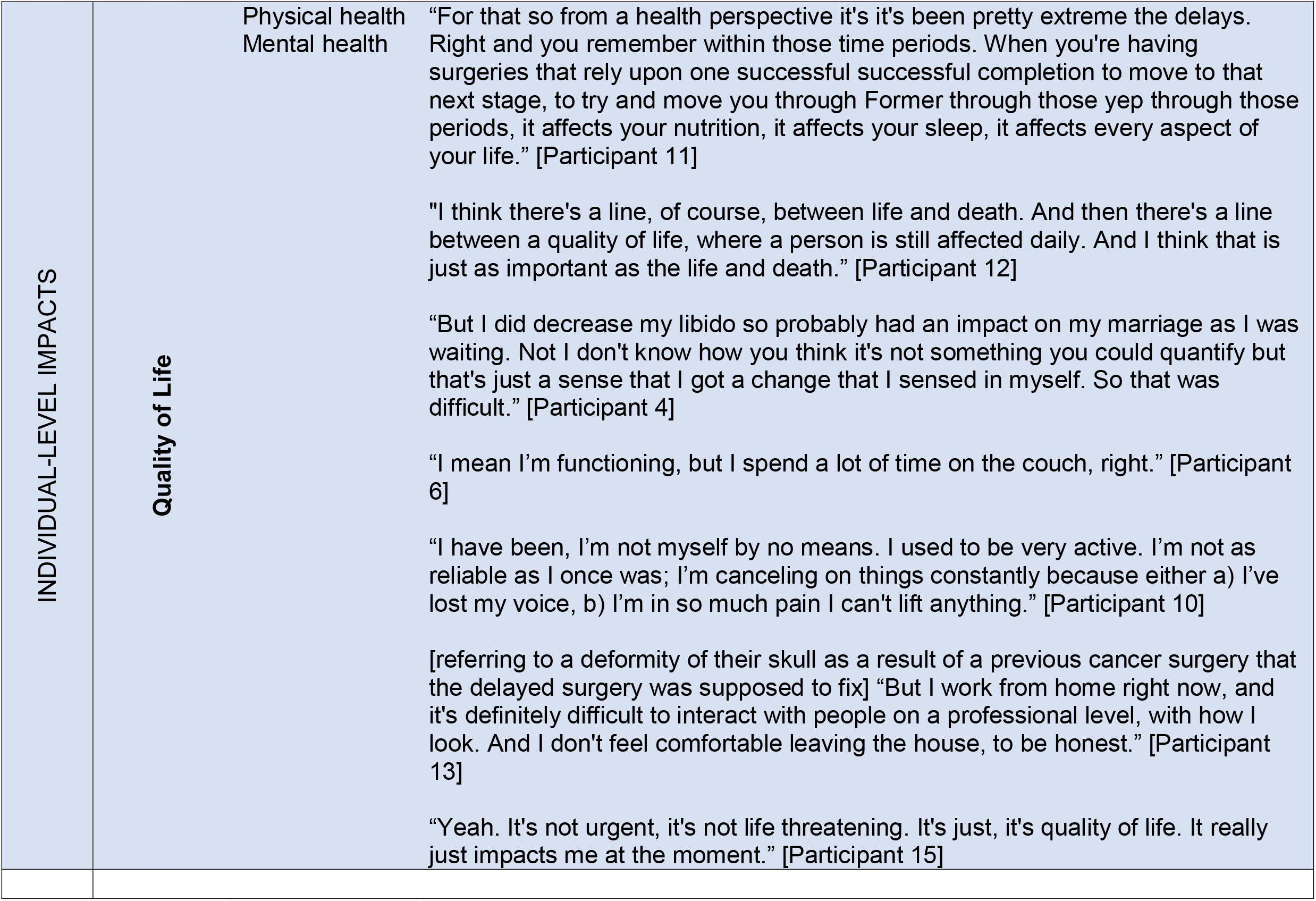
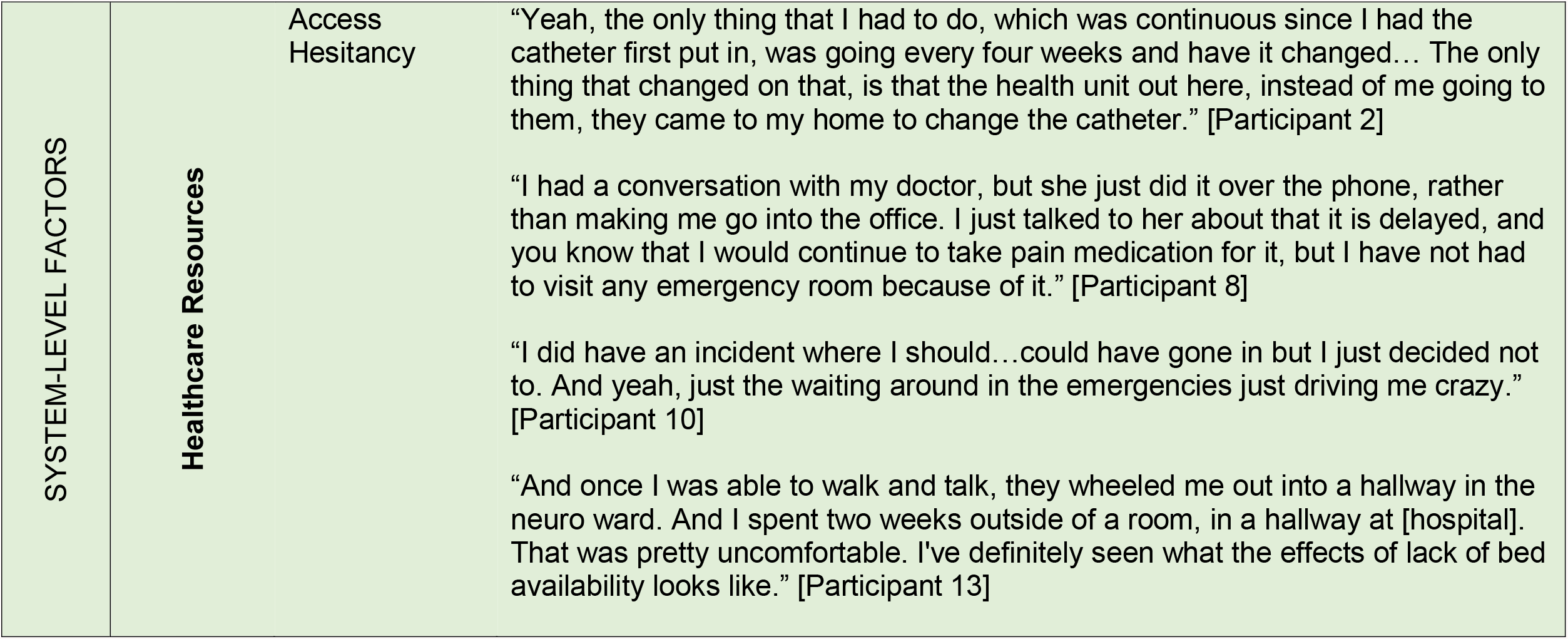
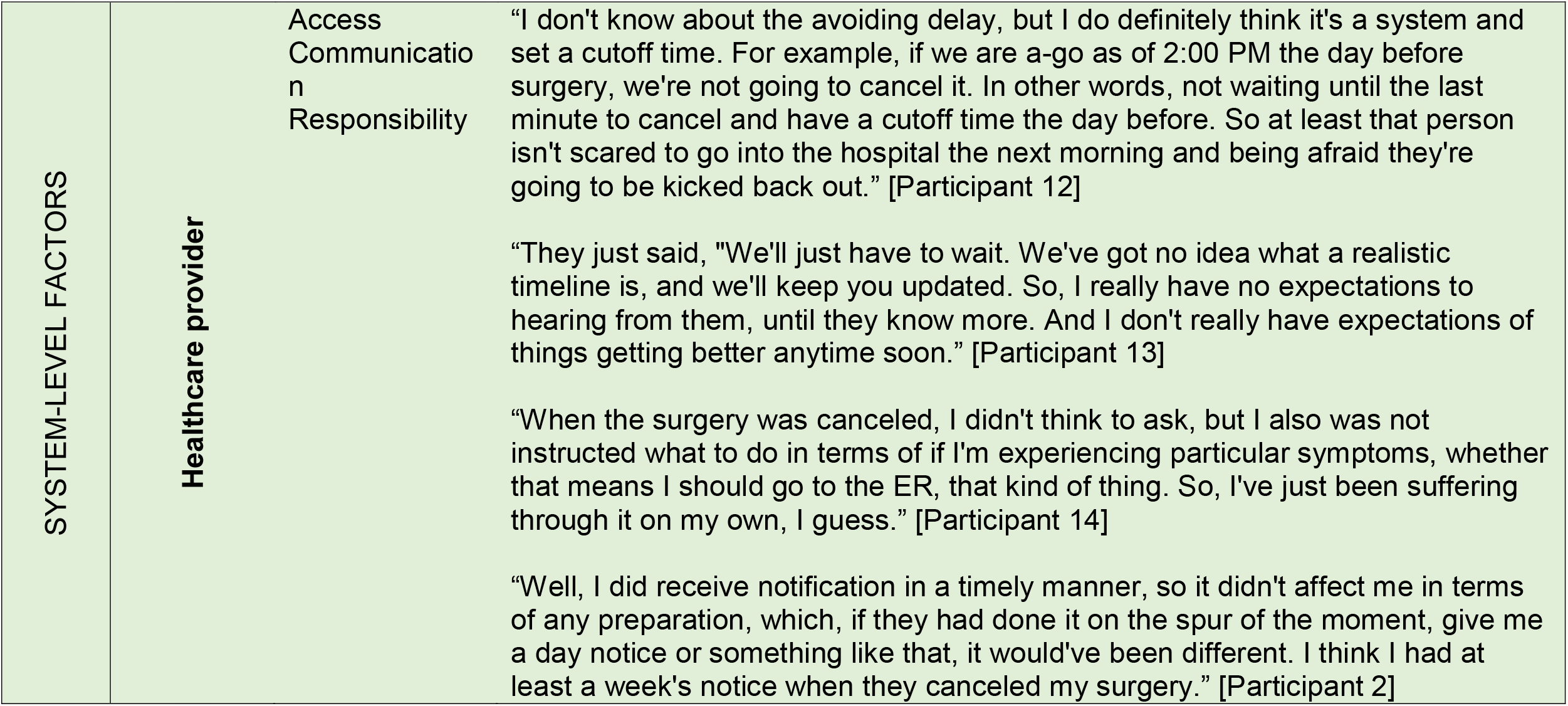
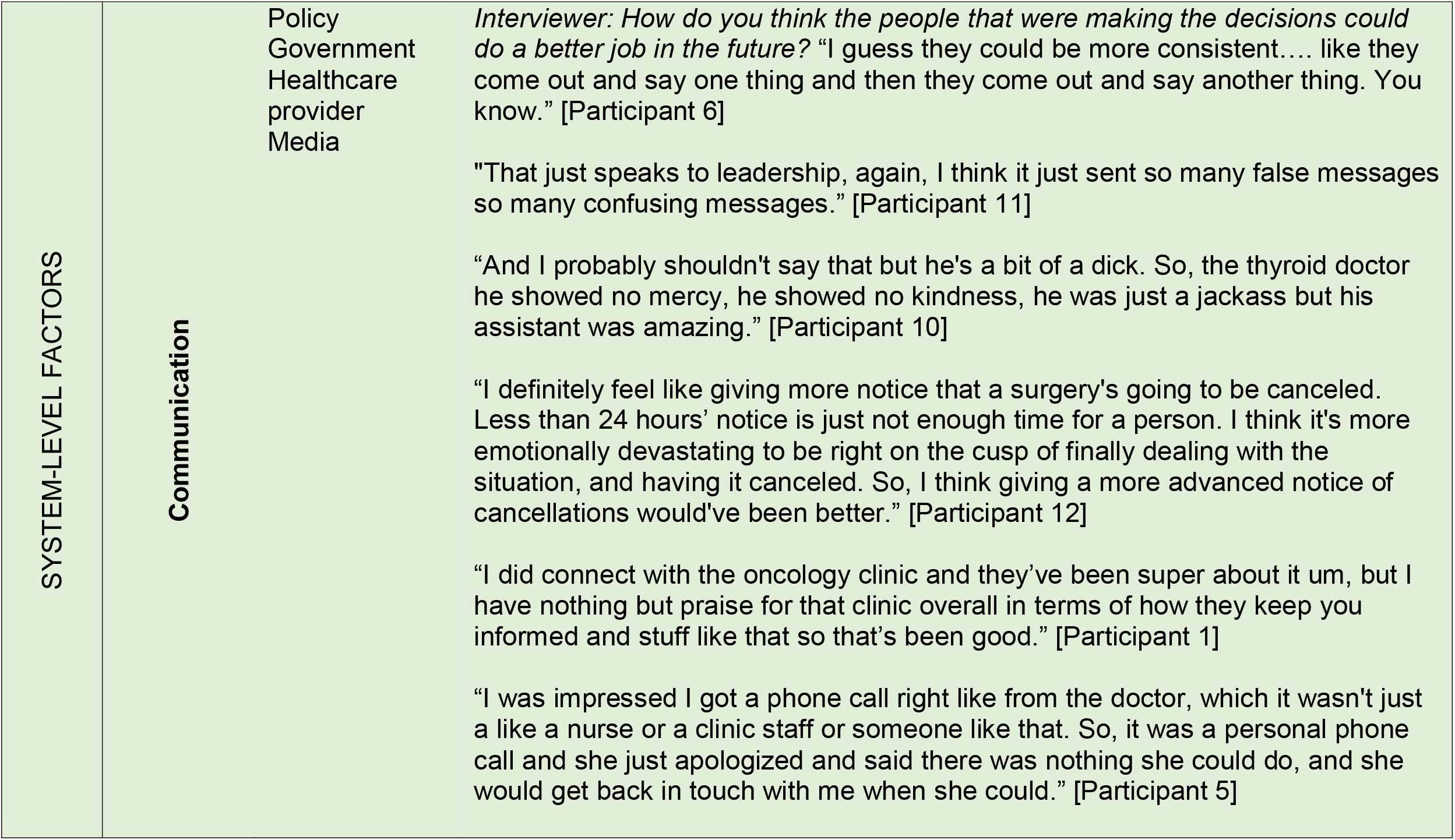
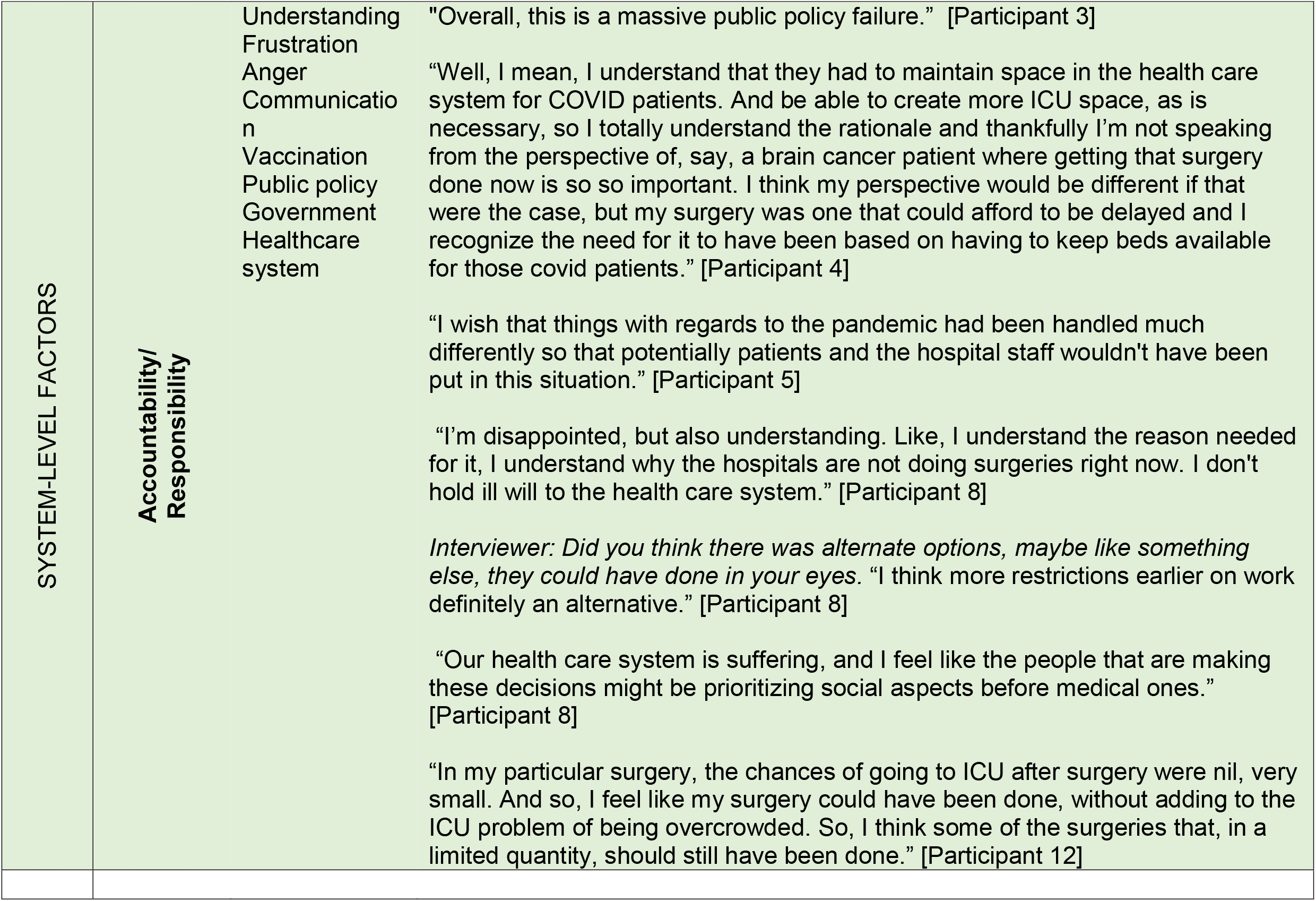
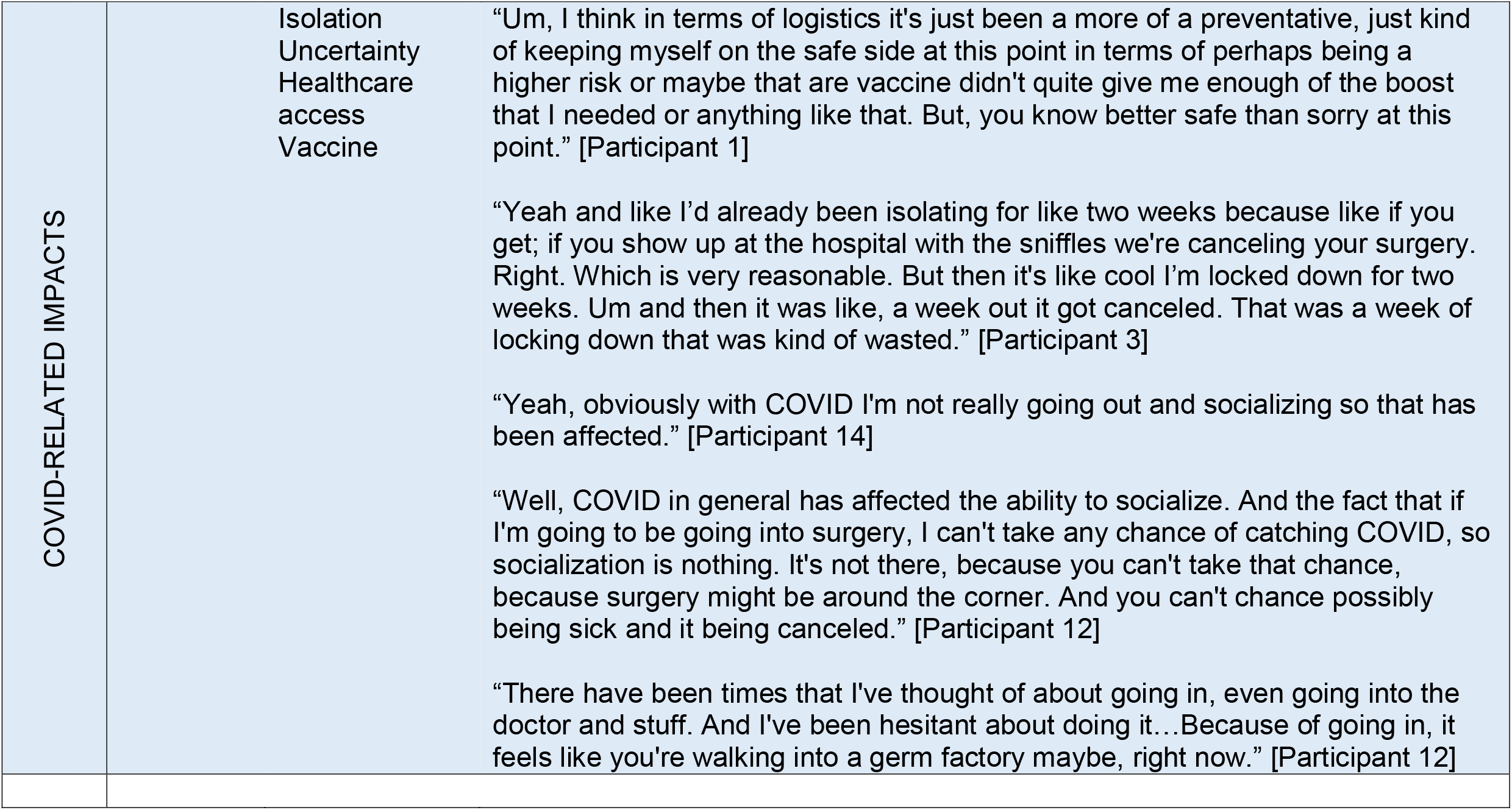
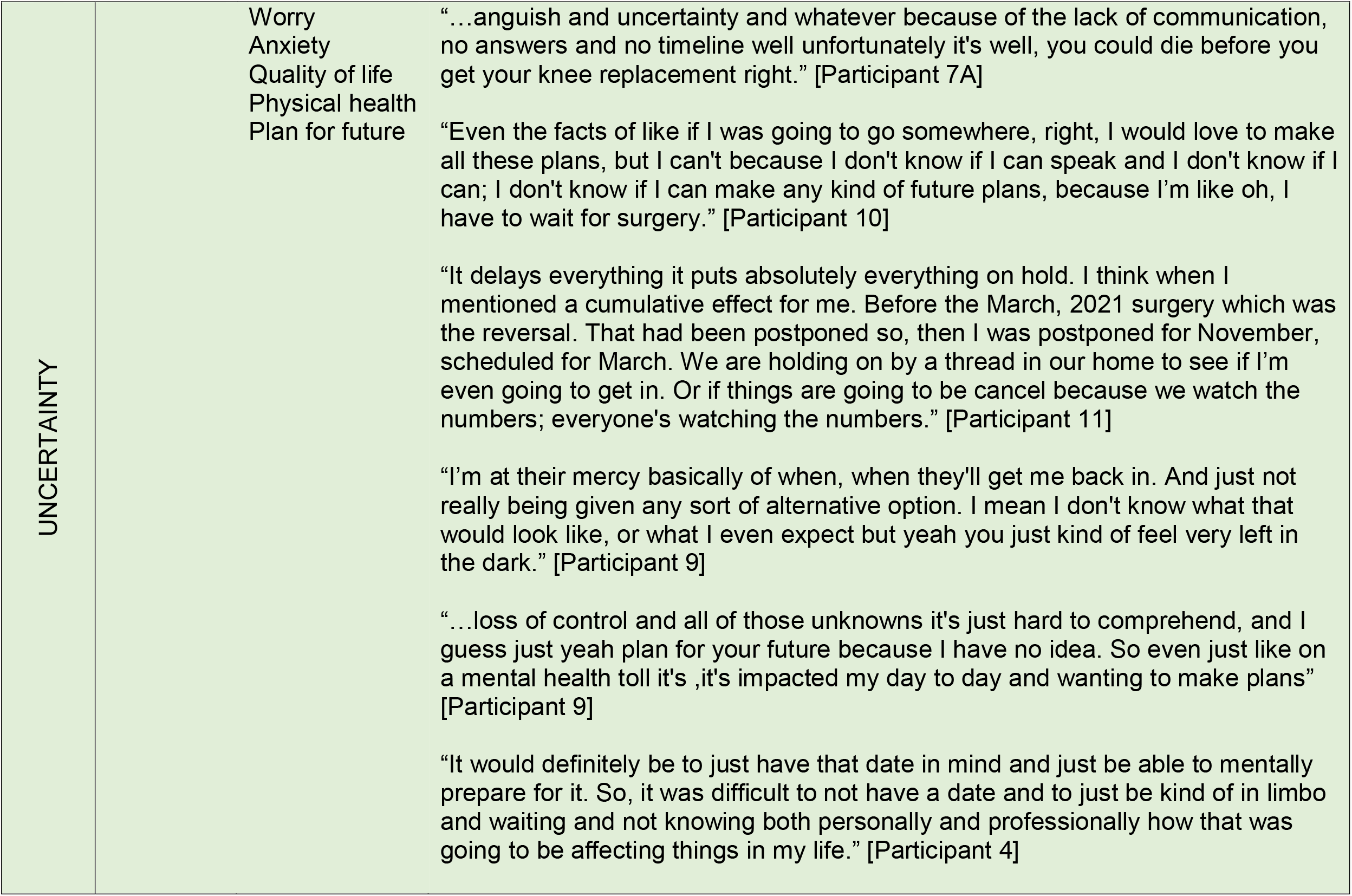
Themes and quotes

#### 1. Individual-level impacts

##### 1. 1. Physical Health

Nearly all participants identified ongoing physical health issues related to their delayed surgery. Physical ailments ranged from mild (“The fibroids in my uterus were getting larger and causing me to become more uncomfortable”) to pervasive and debilitating (“I have a very difficult time breathing, I have a very difficult time eating”). Several patients described ongoing physical pain and the effects on their daily lives; from modifications to exercise and activities of daily living, to inability to do one’s job effectively or at all, and prolonged use of pain medication to manage the pain.

Issues around the impact of delayed surgeries on physical health consequently led to impacts on mental health, specifically around worry and anxiety. Uncertainty related to when the surgery would occur, and relief of physical health consequences was also expressed by many participants.

> “So, this delay, basically I’m afraid I can have a heart attack or stroke, at any time. So, it’s been very stressful on me and my [spouse] as well.”

##### 1.2. Mental Health

All participants described an impact on their mental health and was the most discussed theme. One participant described mental health impacts as their reason for participating in the study:

> “I think the biggest reason why I wanted to participate is; just again, it’s that emotional toll that it takes on people on a daily basis and I have to assume that the government thought about that, but perhaps they don’t realize the actual toll that it takes on all of these other people’s lives…”

The impacts on mental health ranged from disappointment to depression and anxiety. Participants expressed a feeling of depression with the delay of surgery and related to navigating the impacts of the delay. Feeling depressed ranged from passive comments of feeling low, (e.g., “Right now I’m a little depressed… but I keep telling myself let it go”) to newly diagnosed depression. Many participants described anxiety and/or worry that was far reaching and extended beyond physical health (as described above) to concerns about their friends and family, their work and many aspects of their daily life. Many of these feelings of anxiety and/or worry were tied to a loss of control and uncertainty around the plans for their surgery, which are explored in theme four (uncertainty).

##### 1.3. Family & Friends Impact

The impact of delaying surgery extended to family and friends in all cases. This was often related to requiring extra support with activities of daily living (e.g., housework or cooking) or emotional support due to physical and mental health challenges directly related to the delayed surgery. Another commonly described impact was related to social relationships where participants cancelled social plans and trips to visit family and friends out of fear of missing a surgery date. Some participants also expressed safety concerns around contracting COVID or other illness that would compound their existing health issues and prevent them from being able to have surgery.

Several participants also described strains in their relationships outlining that their partners were stressed and worried about the changes in their physical and mental health which in turn caused further stress for patients as they waited for surgery. Some participants also described changes to their sexual relationship with their partners because of difficulty with intercourse because of their unresolved health issues.

##### 1.4. Work Impact

Nearly all participants indicated that their delayed surgery directly impacted their professional life. While some described changes in their ability to work, others disclosed losing their employment; and for one participant this was described as a significant loss to their identity. The theme of uncertainty re-emerged when discussing the impact on the participants’ work life – participants felt that because of the uncertainty around surgical resumption they were unable to plan time off for surgery and this would have a downstream impact on work and colleagues. Similarly, participants held-off taking vacation prior to surgery (prior to the cancellation), which in some cases led to burnout. Other participants noted that they were unable to do their job normally or that they lost employment due to the physical or mental health impacts of waiting for surgery. This led to concerns about job security and many endorsed struggles with earning a stable income and financial stress.

> “I haven’t been able to work as well, which is another factor … I’m going to lose business because of this.”

Conversely, because of flexible, virtual work environments during COVID, some participants highlighted that this allowed them to be able to continue to work while managing their ongoing health issues.

##### 1.5. Quality of Life

For many participants the culmination of impaired physical and/or mental health, strained social interactions, and employment challenges led to poorer quality of life. Interestingly, physical impairments were not always a driving factor; some participants commented that despite being able to physically function, their quality of life still declined. Many patients described their delayed surgery as having an overarching cloud over their lives.

> “At home, I feel like I’m extremely distracted. I’m not present, because I’m constantly thinking about the surgery and the delay. At work, it’s a lack of focus, it’s a worry about how it’s affecting my coworkers. And there’s also the mental exhaustion of not being able to take time off at work because of it.”

#### 2. System-level factors

Participants described several system-level factors primarily revolving around access to and communication with the healthcare system, including their healthcare providers. Furthermore, several participants identified sources of accountability and responsibility for handling of COVID-19 including delayed surgeries and even provided suggestions for how to manage these delays in the future.

##### 2.1. Healthcare resources

Several participants indicated that they required additional care or interacted with the healthcare system, beyond their baseline use, while they were waiting for surgery. This included more frequent visits with their primary care physician (including virtual care visits), consultation with psychologists, regular visits (every four weeks) to change a catheter, regular visits with a gastroenterologist for ileostomy including a day surgery and visits to the emergency department. Some participants expressed less healthcare use due to concerns around contracting COVID-19 in physician offices or hospitals.

##### 2.2. Communication

Discussion around communication focused primarily on communication with their healthcare providers, specifically how and by whom the delay was communicated. Participants’ viewed communication as equally positive and negative overall. Negative comments were largely related to short notice of the delay and lack of information when communicating the delay. Participants who perceived communication to be negative expressed more anger and frustration. Conversely, many participants felt they were given sufficient notice about the delay and praised their healthcare providers for their communication. Participants who reported positive communication indicated that they received timely communication directly from their healthcare provider (surgeon or surgery clinic rather than the healthcare system) who demonstrated regret and concern for the patient.

There were also some concerns raised about communication by the government and media. Specifically, some participants raised concerns that the government and healthcare system communication categorized these surgeries as “elective”; voicing that although their condition may not be immediately life threatening, it was life-altering, and their surgery was far from “elective”.

##### 2.3. Perceived Accountability/Responsibility

When participants were asked what could have been done differently to minimize the impact of the COVID-19 response on surgery patients, discussion frequently deviated to anger and/or frustration about the current situation; specifically, the high number of COVID-19 cases necessitating surge capacity in our hospitals and the resulting decision to delay surgeries.

One participant summarized the overall sentiment of most participants: “I understand it, but I don’t like it”. Anger and frustration with the current situation was frequently associated with uncertainty.

However, other sources of anger and/or frustration were related to responsibility for the COVID-19 situation in our province. Participants frequently cited the government’s handling of the COVID-19 pandemic as the reason for having to delay surgeries. There were also strong feelings of divide towards people who have chosen not to get the vaccine.

> “…surgeries have been delayed, cancer treatments have been delayed. All that with a vaccine on the table. Well, the last thing I checked there wasn’t a vaccine for cancer, so why is someone missing a cancer treatment when the person with COVID who didn’t get immunized.”

Despite negative views about the current situation, including delaying surgeries, there were also sentiments of understanding, especially regarding the decision to delay surgeries during the first wave of COVID-19 when less was known about the trajectory of COVID-19 and when vaccines were not available.

#### 3. COVID-19-related impacts

Participants highlighted unique factors related to COVID-19 that complicated their delayed surgery experience. Protecting and maintaining one’s health and isolation were two common threads around COVID-19 impacts.

Participants expressed concern or challenges with accessing healthcare services due to COVID-19 but there were also some advantages to accessing care during COVID-19 with the advent of virtual care. Some participants indicated hesitation in accessing in-person healthcare for fear of acquiring COVID-19 in public places. Similarly, the fear of contracting COVID-19 or other illnesses in their vulnerable condition was a great concern. The fear was twofold; they were fearful of becoming severely ill but also fearful of further surgical delays if they were to become ill at a time when a surgery date became available. Their fear of becoming ill resulted in physical isolation and consequently psychological isolation for many participants. Moreover, their poor mental health also made socialization (virtual or in-person) more challenging.

#### 4. Uncertainty

Uncertainty about health and surgery resumption had a large impact on participants. All participants found the uncertainty surrounding their surgery and physical health was pervasive and impacted many aspects of their life. Uncertainty was described as a loss of control and lack of information about surgery and underlying health. One participant described this loss of control as “being held hostage to what’s happening in the hospital; like…total limbo”.

In addition to feelings of uncertainty resulting from the perception of loss of control and being uninformed, uncertainty was also described as a reason for poor mental health. This association was rooted in feelings of anxiety and/or worry about the unknown consequences of delaying surgery on their physical health and uncertainty about the future.

Uncertainty resulted in poor mental health but also logistic challenges in several areas of life. For example, uncertainty impacted planning time off work for surgery and the subsequent recovery. As well, it impacted the participant’s ability to commit to social activities, engage in family planning and pursue post-secondary education. Uncertainty frequently led to discussion about the impact of the delayed surgery on friends and family and their professional lives.

### Interpretation

Our study found that the impact of delaying surgeries to manage the surge of COVID-19 patients was diffuse and consequential on the lives of patients. Four interconnected themes emerged: 1] individual-level impacts (subthemes: physical health, mental health, family and friend impact, work impact, quality of life), 2] system-level factors (subthemes: communication, healthcare resources, perceived accountability/responsibility), 3] COVID-19-related impacts and 4] uncertainty (Figure 2).

The physical effect of delaying surgery has been illustrated in the pre-pandemic setting. Exceeding optimal interval from diagnosis to surgery has been associated with poor outcomes in many diseases.^(7, 8, 15-17)^ For example, delayed cancer surgery results in poor disease-free survival,^(17-19)^ and patients with delayed cardiac surgery experience greater morbidity and mortality, longer hospitalizations, and are more likely to die post-operatively.^(20, 21)^ Delaying surgeries considered less urgent, is associated with an increase cost to patients (e.g., increased pain, decreased quality of life) and the healthcare system.^(22-26)^ Our study confirmed that perceived physical health was compromised due to the delays in surgery; however, the physical impact was less prominent and perhaps the driver of more significant effects on participants’ mental health and quality of life.

Distress among patients waiting for surgery has been previously described, however there is a need for additional research on the burden of delayed surgeries during COVID-19 given the unique and unprecedented nature of surgical delays during the pandemic.^(27)^ A recent study of patients waiting for cancer surgery due to COVID-19 capacity limits reported similar findings to ours; participants had high levels of distress, stress, anxiety and depression.^(9)^ The similarity between our findings and those of Forner et al.,^(9)^ despite few cancer surgery participants in our study suggests the experience of waiting for surgery during COVID-19 is similar across surgeries. We hypothesize the unifying characteristic among patients waiting for any type of surgery during COVID-19 is uncertainty; specifically, the unpredictability of the COVID-19 pandemic and response. Uncertainty was a prominent theme that arose in our study and many participants underlined the connection between the uncertainty and the distress and burden this placed on all aspects of their lives. Minimizing uncertainty is challenging in the context of COVID-19 because the pandemic has been, in and of itself, unpredictable. Our findings highlight that timely, personalized, pragmatic, and compassionate communication is needed and may minimize distress in patients waiting for surgery, even if certainty cannot be provided.^(27-29)^

#### Moving forward

Managing surgical waitlists and transforming surgical care is important to mitigate the backlog of surgeries due to the response to COVID-19. Approaches to decrease wait time and increase efficiency of surgical care to create more capacity, and increase quality, have been proposed.^(30-32)^ However, focus on improving the surgery waiting period could minimize the impact of delaying surgery during and after the COVID-19 pandemic. The waiting period is fraught with consequences to patients’ physical and mental health.^(7)^ Based on our findings, interventions that support patient mental health are needed during surgery waiting periods. Studies suggest that strategies such as educating patients about the potential mental health consequences of waiting for surgery, regular communication with healthcare providers and mental health support (e.g., self-management approaches and peer-support) could improve the impact of surgical delays on patients’ mental health.^(27)^ Our findings support the need and potential utility of these strategies but future research needed to explore the critical components for successful strategies and their implementation.

#### Strengths and limitations

While this study has several strengths (data collection continued after we reached thematic saturation to ensure a broad perspective of a varied patient sample, we used iterative member checking, we captured patient perspectives from diverse backgrounds and clinical areas and our team engaged in peer debriefing during analysis) our findings should be interpreted with limitations in mind. Participants were limited to a single province, during the largest wave of the COVID-19 which may differ from the unique experience with COVID-19 of each province.

## Conclusion

During the ongoing COVID-19 pandemic, healthcare systems have had to make difficult decisions with little evidence. While healthcare system decision-makers anticipated the impact of delaying surgeries on patients, they had little choice due to strained healthcare resources. Our findings illustrate the direct and substantial impact delaying surgery has on patients. The interplay between the themes identified in our study suggests it is important to consider all these factors when measuring and developing strategies to mitigate the impact of delaying surgery. Investments in mental health, occupational and social supports with clear, personalized, and compassionate communication strategies during periods of delay could address all these factors and should be explored to improve surgical care during the ongoing COVID-19 pandemic and beyond.

## Supporting information

Semi-structured interview guide

COREQ checklist

## Data Availability

All data produced in the present study are available upon reasonable request to the authors

## Notes

**Competing Interests:** Authors have no conflicts of interest to declare.

### Competing Interest Statement

The authors have declared no competing interest.

### Funding Statement

This study did not receive any funding

### Author Declarations

This study was approved by the University of Calgary Conjoint Research Ethics Board (REB20-0753).

