## Supplementary material for "“It affects every aspect of your life”: A qualitative study of the impact of delaying surgery during COVID-19": Semi-structured interview guide

Thank-you for taking the time to participate in this interview to help us understand the impact of delaying non-urgent surgeries on patients. My name is {insert name}, I am a {insert role}. The primary investigator is Dr. Khara Sauro.

You have been invited here today because we are interested in hearing your experience as a patient who had a planned surgery delayed during the COVID-19 pandemic response. We will ask for your experience and ask a few questions about the type of surgery you were planning to have, how that affected your health and everyday life.

Thank you again for your participation.

Before we begin, I would like to make sure that you know that there are no right or wrong answers in our discussion today; we are interested in all comments. We don’t want to miss any of your comments and feedback, so we are taking detailed notes and audio recording our conversation.

Please be assured what is said and discussed today will be kept confidential and any information you provide will be kept in a password protected folder on a secure server, accessible only by members of the research team. As is noted in the informed consent, your participation is voluntary and you are free to decline participation at any point should you wish to do so for any reason. Please let me know if you have any questions.

Do you have any questions before we begin?

1. Please describe the surgery you were booked to have, as you understand it.
2. When were you scheduled to have your surgery and do you have a new date for your surgery?
3. Is this the first time your surgery is being re-scheduled?
4. Can you please tell us about how having your surgery delayed has affected your overall health?

Prompts:

a. How has your healthy been since your surgery was delayed?
b. What about your mental health?
c. How have you interacted with your healthcare provider(s)? Is that different than before you were supposed to have surgery?
d. Have you had to visit the emergency room or hospital while you were waiting for your surgery? Can you tell me about that?
5. Can you please tell us about how having your surgery delayed has affected your everyday life, such as activities of daily living, work, income, socialization, family, relationships?

*Ask this only if they haven’t touched on this question in their response to the previous question.

Prompts:

a. Have you had to change many aspects of your daily living because your surgery was delayed? Can you tell me more about that?
b. Have you had to ask for more help from family or a caregiver? Can you expand on that?
c. Have you been able to do the things you normally would do?

What do you think about the decision to delay surgeries?

Prompts:

a. Was the right thing to do given the circumstances?
b. What do you think the alternate options were?
c. How could the people making the decisions do a better job in the future?
d. What do you think was done really well?

I would just like to take a moment and summarize what we’ve talked about today. <Summarize key points>.

Do you have anything else to add that we may not have captured?

Thank you so much for speaking with me today and for providing your experiences.
